# Topical Insulin as a Novel Treatment for Persistent Epithelial Defects and Other Ocular Surface Disorders: A Systematic Review

**DOI:** 10.1101/2024.10.25.24316042

**Authors:** Raluca Bievel Radulescu, Stefano Ferrari, Diego Ponzin

## Abstract

**Purpose:** Through this systematic review, we evaluated the therapeutic potential of topical eye insulin in different concentrations to treat several surface ocular pathologies, including: persistent epithelial defects, diabetic keratopathy after a vitrectomy, neutrophic keratopathy and dry eye syndrome. We have consolidated through the data, what are the doses used, the methods of preparation for insulin, whether there are adverse effects and what would be the effectiveness of the eye drops with insulin.

**Methods:** We carried out an extensive search including Pubmed, Cochrane Library, Scopus and Web Of Science. We found 43 relevant studies, after which we excluded duplicates, animal studies, case reports, we ended up with 14 studies to include in the article. Through the Newcastle-Ottawa scale for observational studies and the Jadad scale for randomized controlled trials, we investigated the methodological quality of these articles.

**Results:** Within the review we included a significant number of 525 patients who used eye drops with insulin in concentrations from 0.5 to 2 U/ml, having an ocular benefit in corneal healing rates without adverse effects. The quality analysis of the included studies showed a NOS score of moderate-high quality, whereas the Jadad scale showed a high quality.

**Conclusions:** Our systematic review demonstrates that patients with persistent epithelial defects, diabetic keratopathy following vitrectomy, neurotrophic keratopathy, and dry eye syndrome showed significant improvements in corneal healing rates. To gain a clearer understanding of the effectiveness of insulin eye drops, future research should include direct comparisons with autologous serum eye drops and amniotic membrane eye drops. These studies will help establish comprehensive clinical guidelines.

## Introduction

Despite all the advances in ophthalmic care, several of these surface disorders are challenging and may present with significant visual impairments that complicate their management. The spectrum may involve persistent epithelial defects (PEDs), diabetic keratopathy (DK) post-vitrectomy, neutrotrophic keratopathy (NK), and dry eye syndrome (DED). In general, these are caused by specific underlying causes such as limbal stem cell deficiency, diabetes, trauma, inflammation, or infection leading to impaired corneal healing with loss of ocular surface integrity. The standard treatments of lubricating eye drops, autologous serum, and surgical interventions usually succeed little in the refractory state of the disease, so other modes of therapy need to be considered (1–3). While insulin is best known for its role in the regulation of blood sugar levels, it actually produces a series of metabolic and cellular activities, thus being a promising candidate to induce corneal healing with the anabolic effects it produces.

Recent studies have highlighted the therapeutic role of insulin in the healing of the ocular surface. Receptors for insulin and IGF, on the surface of the cornea-keratocytes and epithelial cells, are able to mediate the migration of cells and influence growth and developmental regulation in the cells of the corneal epithelium, an essential process for renewal of the corneal surface (4). Diabetic animal models have been subjected to several experimental studies. Insulin eye drops have been shown to normalize DNA synthesis in basal epithelial cells within 48 h from injury. These data suggest that insulin may increase cell proliferation and thereby promote the process of reepithelialization (5)This mechanism is relevant in conditions where corneal epithelial healing is impaired, providing a possible explanation for the beneficial effects of topical insulin obtained in clinical settings. Insulin-like growth factors, especially IGF-1, may play a central role in the growth, differentiation, and proliferation of corneal epithelial cells. (6,7)

These receptors and insulin receptors are both present on the surface of corneal keratocytes and epithelial cells, evidencing a very strong biological relationship between these growth factors and corneal integrity. Insulin-a similarly powerful anabolic hormone which is structurally related to the IGFs-has been found to be present in tear film with a mean concentration of 0.404 ± 0.129 ng/mL (7) and thus is thought to affect processes of corneal healing. These findings have outlined the need for a structured study to analyze the power of eye drops with insulin in the treatment of different ocular surface pathologies, focusing their attention on persistent epithelial defects, diabetic keratopathy after vitrectomy, neutrotrophic keratopathy, dry eye syndrome.

This review aims to outline the current research on topical insulin therapy in ophthalmology, assessing for efficacy and safety while highlighting areas of future research needs. This review will synthesize findings from a number of studies in a manner that will be informative to clinical practices and supportive in the integration of topical insulin into protocols in the management of complex ocular surface disorders.

## Methods

### Systematic Review Protocol

This systematic review was conducted by using the PRISMA (Preferred Reporting Items for Systematic Reviews and Meta-Analyses) statement and supported the Meta-analysis of Observational Studies in Epidemiology (MOOSE) guidelines for systematic reviews of observational studies.

### Identification of Studies

An extensive literature search was made by use of different electronic databases: Pubmed, Cochrane Library, Scopus and Web Of Science to identify the studies related to the use of topical insulin eye drops in the treatment of ocular surface disorders. Various combinations of keywords used included "topical insulin," "ocular surface disease", "persistent epithelial defects", "diabetic keratopathy", "neurotrophic keratopathy," "dry eye disease", "corneal healing" and "insulin eye drops." No date of publication was restricted, and the search was therefore made for all literature available up to August 2024.

The references included in the selected papers were also checked to see if other studies could have been missed while searching in the database. During the selection phase, first-step selection of titles and abstracts was performed by the primary reviewer, followed by full-text review of all those articles that appeared relevant. To validate the process of inclusion, a second independent rater reiterated the whole selection process. Any disagreement on study inclusion or exclusion was discussed to a consensus, and an arbitrator was consulted when there was a need for that. The methodological quality of each included study in the review was assessed using appropriate tools depending on the study design. In the case of randomized controlled trials, methodological quality was assessed by means of the modified Jadad scale, which assesses items related to randomization, blinding, and withdrawals/dropouts. For observational studies, the Newcastle-Ottawa Scale was applied. The data extracted from each article included study design details, the country where the study was performed, the characteristics of participants, insulin concentration, and administration regimen, outcome measures, potential confounders, key findings of the study.

Studies were included in the review based on the following criteria:

1. The study was conducted to examine the role of topical insulin eye drops in ocular surface disorders including but not limited to persistent epithelial defects (PEDs), diabetic keratopathy (DK), neurotrophic keratopathy (NK), and dry eye disease (DED).
2. Studies that provided data on the efficacy, safety, or both, of topical insulin in promoting corneal epithelial healing.
3. Studies including human subjects irrespective of age or condition.
4. Full-text published works.

Exclusion criteria were as follows:

1. Case reports, abstracts, letters, and editorials.
2. Animal model studies and in vitro experiments.
3. Articles dealing exclusively with the biochemical properties of insulin without including data on clinical use.

### Data Extraction

The titles and abstracts of the eligible articles were screened for relevance. Full-text articles were further assessed, whereby the two independent reviewers extracted data on the following: 1. Study design: Type of study- e.g., randomized controlled trial, retrospective study, prospective study, case-control study; 2. Objectives: The primary aim of the study related to the use of topical insulin; Population: Characteristics of participants, including ocular conditions treated and sample size.

4. Intervention: Details of insulin concentration, dose, and frequency of application.

5. Outcome Measures: Primary and secondary outcomes pertaining to rates of healing of the cornea, changes in staining of the cornea, tear break-up time, amongst other such relevant measures of clinical interest.

6. Safety Profile: Any adverse effects or concerns for safety regarding topical insulin use.

7. Results: Statistical significance of main findings, effect size, and comparisons with control or other treatments.

8. Confounding: Evaluation of confounders and their adjustments in the studies

9. Limitations regarding the study: Also comment on the limitations and biases reported in the reviewed studies.

Lastly, we evaluated the methodological quality of each eligible study included using appropriate tools such as the Newcastle-Ottawa Scale for observational studies to ensure the reliability and validity of the results.

## Data Synthesis

Results from the selected literature were summarized to give an overview of topical insulin’s efficacy and safety as a therapeutic agent for ocular surface disorders. The results of studies included were summarized narratively, and where appropriate, a meta-analysis was considered by combining the findings quantitatively. Where heterogeneity existed in study design, populations, or outcomes, these were discussed to contextualize the results.

## Data Extraction

The titles and abstracts of the eligible articles were screened for relevance. Full-text articles were further assessed, whereby the two independent reviewers extracted data on the following:

1. **Study design:** Type of study- e.g., randomized controlled trial, retrospective study, prospective study, case-control study
2. **Objectives:** The primary aim of the study related to the use of topical insulin
3. **Population:** Characteristics of participants, including ocular conditions treated and sample size.
4. **Intervention:** Details of insulin concentration, dose, and frequency of application.
5. **Outcome Measures:** Primary and secondary outcomes pertaining to rates of healing of the cornea, changes in staining of the cornea, tear break-up time, amongst other such relevant measures of clinical interest.
6. **Safety Profile:** Any adverse effects or concerns for safety regarding topical insulin use.
7. **Results:** Statistical significance of main findings, effect size, and comparisons with control or other treatments.
8. **Confounding:** Evaluation of confounders and their adjustments in the studies
9. **Study limitations:** Reported limitations and biases within the studies.

Lastly, we evaluated the methodological quality of each eligible study included using appropriate tools such as the Newcastle-Ottawa Scale for observational studies to ensure the reliability and validity of the results.

## Data Synthesis

Results from the selected literature were summarized to give an overview of topical insulin’s efficacy and safety as a therapeutic agent for ocular surface disorders. The results of studies included were summarized narratively, and where appropriate, a meta-analysis was considered by combining the findings quantitatively. Where heterogeneity existed in study design, populations, or outcomes, these were discussed to contextualize the results.

## Results

A total of 43 studies were identified from the database searches, of which 9 duplicates were excluded. After reviewing the titles and abstracts, 13 studies were discarded because they did not conform to the inclusion criteria; of the remaining 21 a further 14 were discarded (inappropriate populations, missing data, etc.) leaving 14 papers, the full manuscripts of which were assessed for eligibility. (Figure 1)

**Fig. 1.**
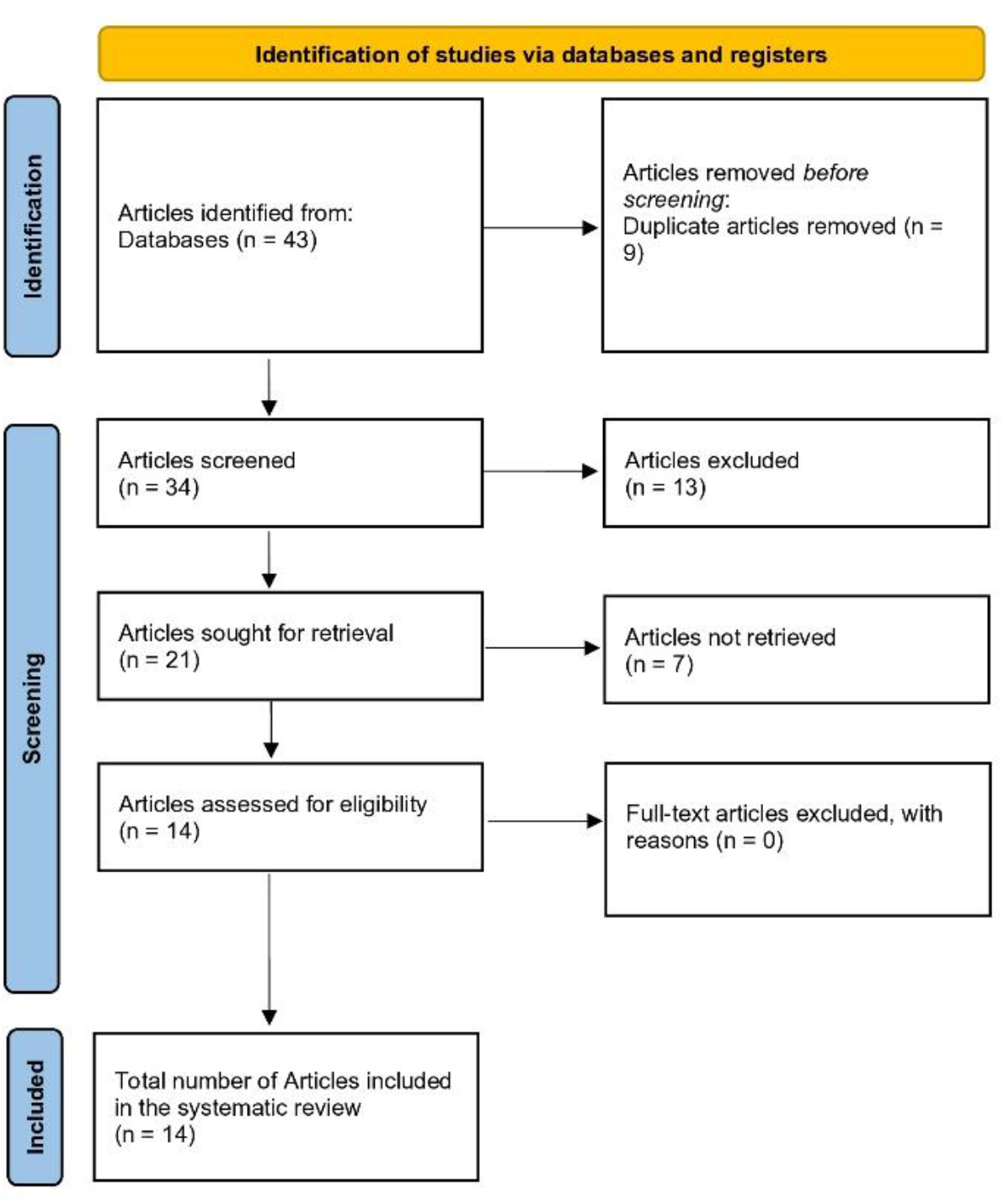
PRISMA flow diagram of the study selection process with reasons for inclusion and exclusion

The Newcastle-Ottawa Scale was applied for the assessment of methodological quality for observational studies included and the Jadad scale for randomized trials. Tables 1 and 2 depict the itemized scores for each study. Observational studies included were independently assessed by two researchers, with an average score of 6.71; thus, a moderate-high quality was detected in the studies. The mean Jadad score from the randomized trials was 7.5 points, indicating high quality study.

**Table 1.**
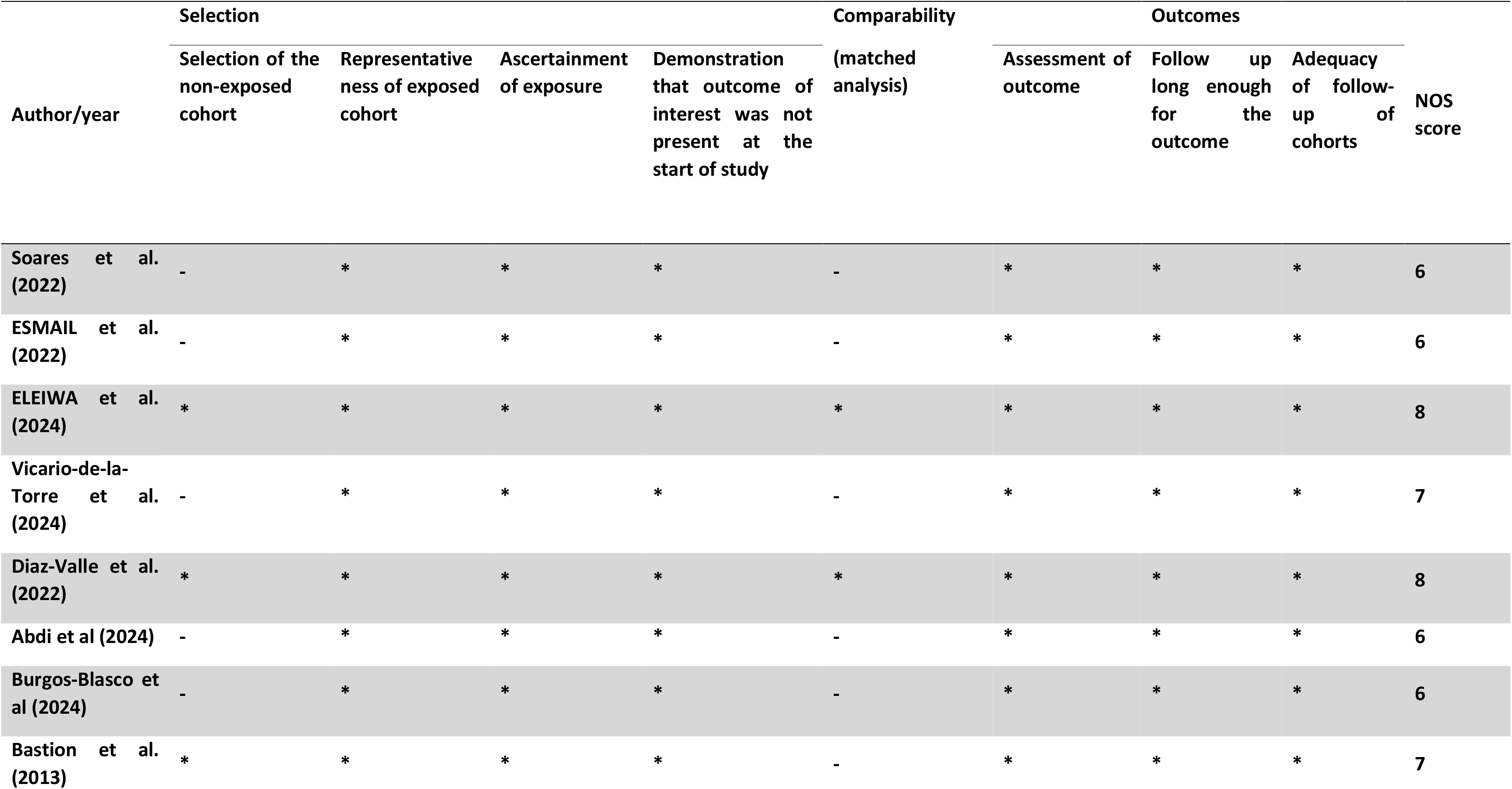
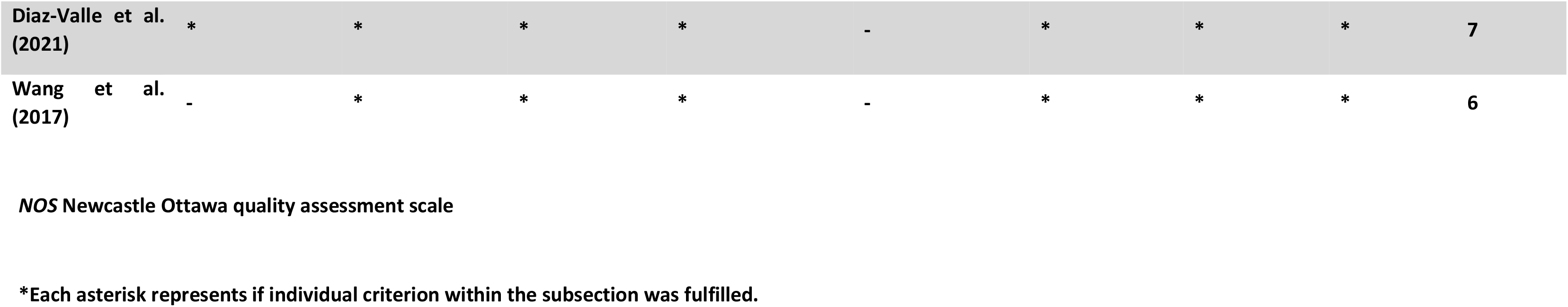
Study quality assessment using Newcastle-Ottawa scale for observational studies.

**Table 2.** Quality assessment of included trials using the Jadad scale.

| **Author/year** | **Was the research described as randomized** | **Was the approach of randomization appropriate** | **Was the research described as blinding** | **Was the approach of blinding appropriate** | **Was there a presentation of withdrawals and dropouts** | **Was there a presentation of the inclusion/exclusion criteria** | **Was the approach used to assess adverse effects described** | **Was the approach of statistical analysis described** | **Total** |
| --- | --- | --- | --- | --- | --- | --- | --- | --- | --- |
| **Quiroz-Mendoza et al. (2021)** | **1** | **1** | **0** | **0** | **1** | **1** | **1** | **1** | **6** |
| **Fai et al. (2017)** | **1** | **1** | **1** | **1** | **1** | **1** | **1** | **1** | **8** |
| **Dasrilsyah et al. (2022)** | **1** | **1** | **1** | **1** | **1** | **1** | **1** | **1** | **8** |
| **Azmi et al. (2020)** | **1** | **1** | **1** | **1** | **0** | **1** | **1** | **1** | **7** |

### Study characteristics

This systematic review encompasses 14 studies, including 5 randomized controlled trials, 4 retrospective studies, 3 prospective studies, and 2 case-control studies. A total of 525 participants are represented in these studies. The application of topical insulin eye drops in the treatment of PEDs, DK, NK, DED, and other ocular surface disorders is dealt with in these studies.

The topical insulin concentrations explored ranged from 0.5 to 2 units/mL and were mostly administered four times daily. The articles ranged in date from the year 2013 to 2024. Among patient groups, there were five studies involving diabetic patients, three involving neurotrophic keratopathy of different etiologies, three involving PED patients, and two other patient populations with dry eye disease.

Some of the measured outcomes that varied in the studies included the rate of resolution of defect epithelium, change in corneal staining, tear break-up time, Schirmer test score, and rate of healing of a corneal epithelial wound. In addition, all had been sourced from different locations such as Iran, Spain, Malaysia, Brazil, and many other countries for a wide demographic representation of the participants.

Across the studies, there are consistent findings that the topical insulin eye drops have been effective and safe for various disorders of the ocular surface, noting scant adverse effects. Some studies indicated marked improvements in the corneal healing rate and reductions in corneal staining, along with overall improvement of ocular surface health, thereby underpinning the potential of topical insulin as a promising alternative therapy.

Table 3 summarizes all the selected studies that have demonstrated topical insulin administration associated with improved clinical outcomes in the management of ocular surface disease. Accordingly, short discussions on findings and individual characteristics of each study are presented herewith for better understanding of the role of insulin eye drops in ocular surface disorders.

**Tabel 3.**
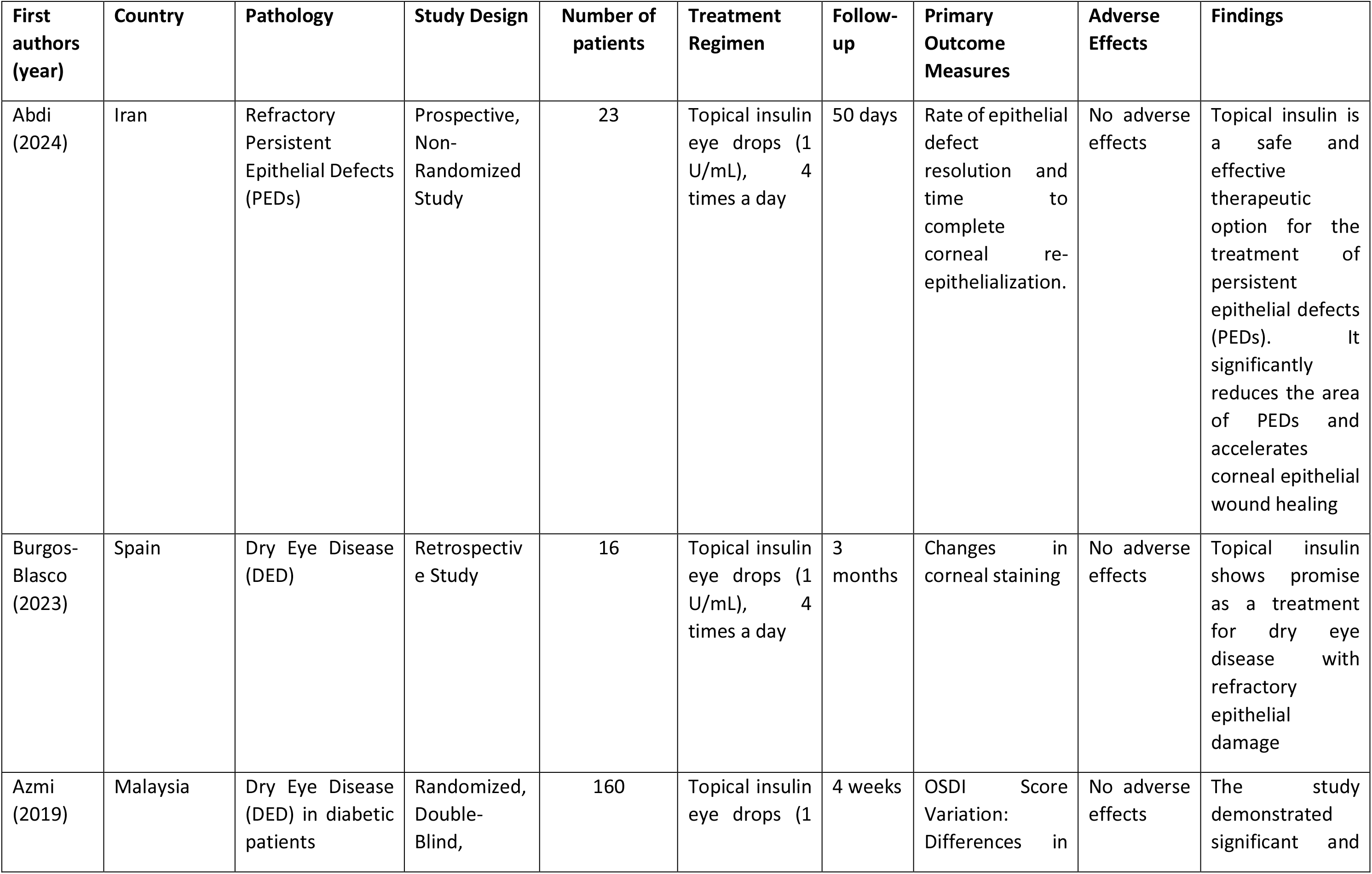
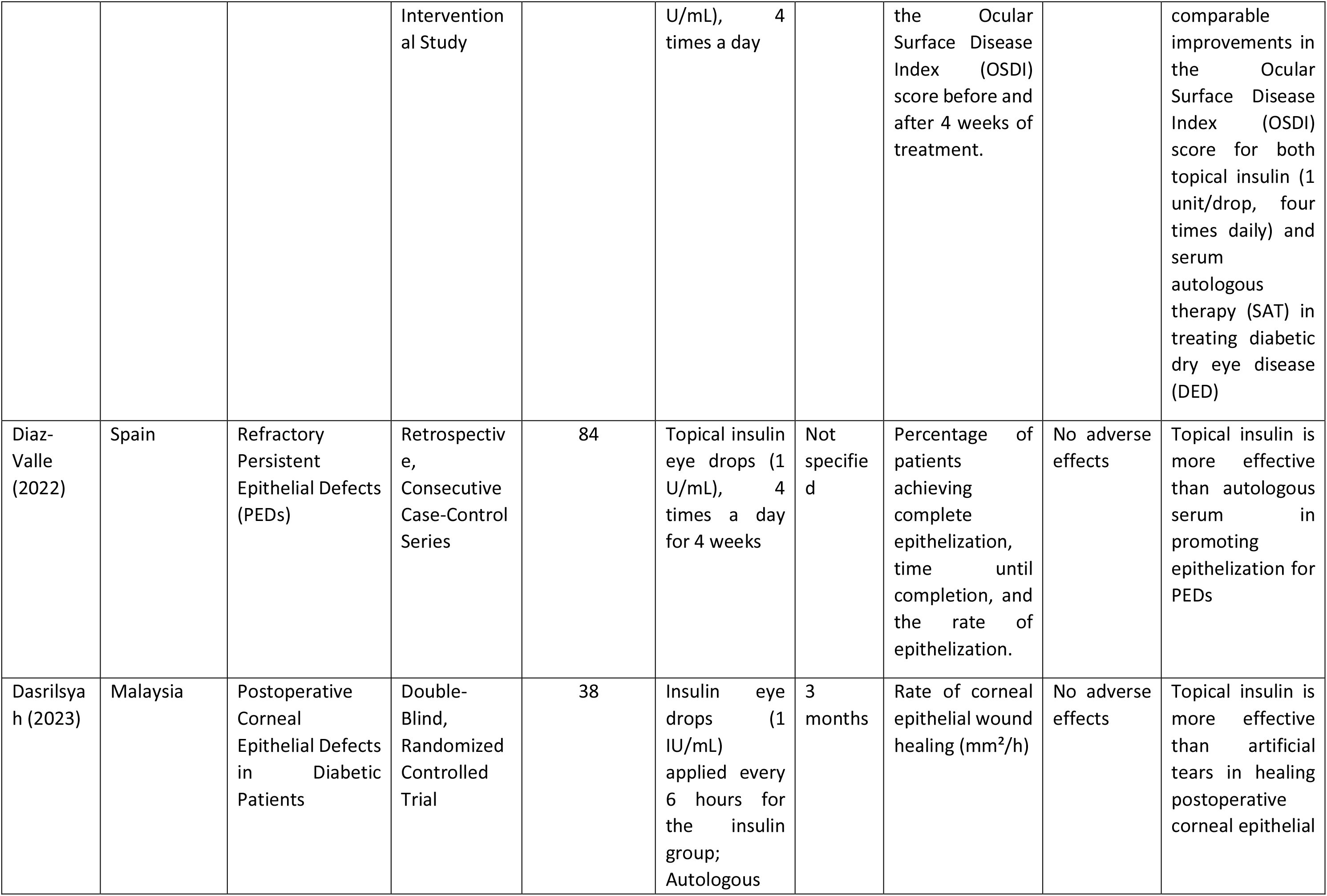
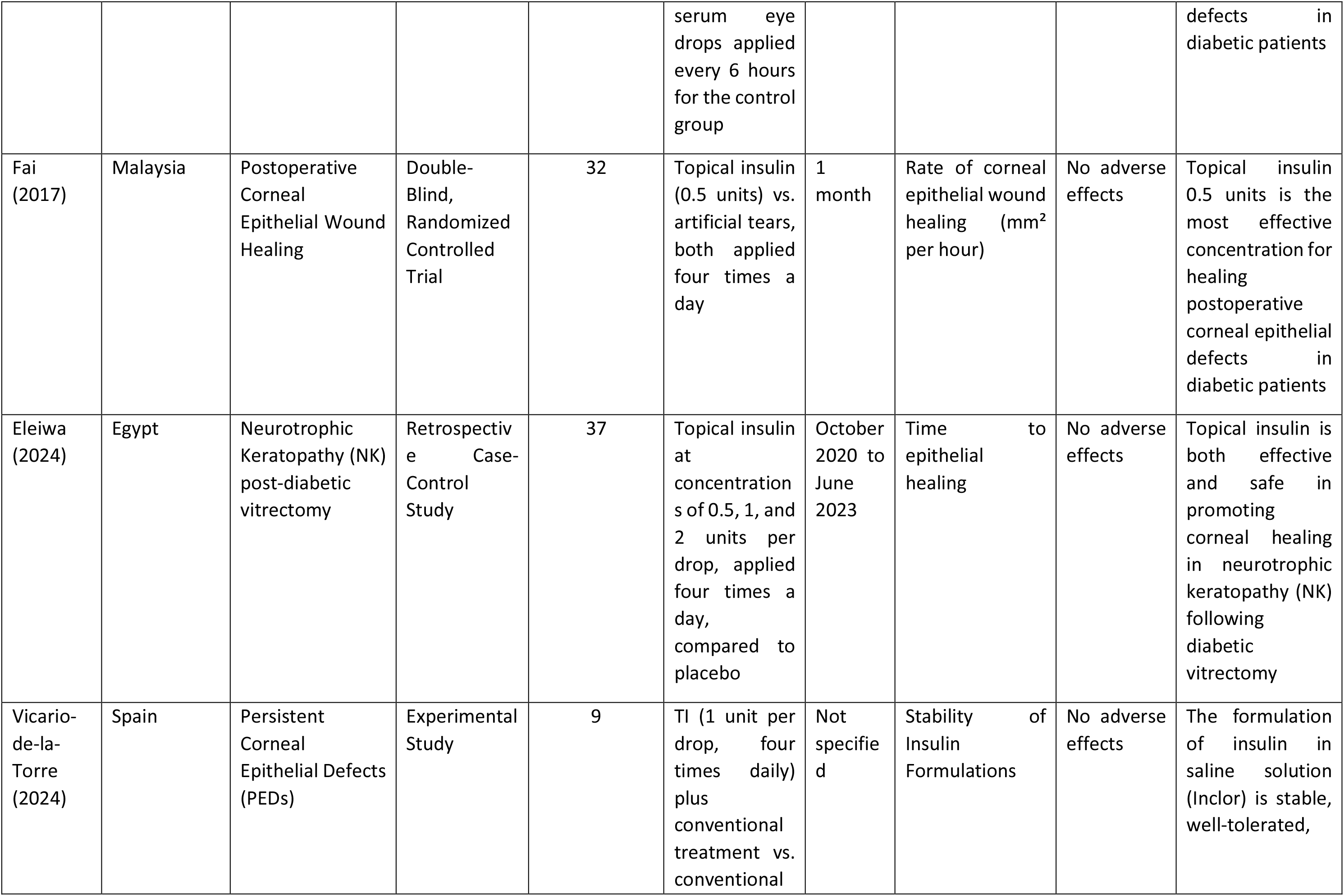
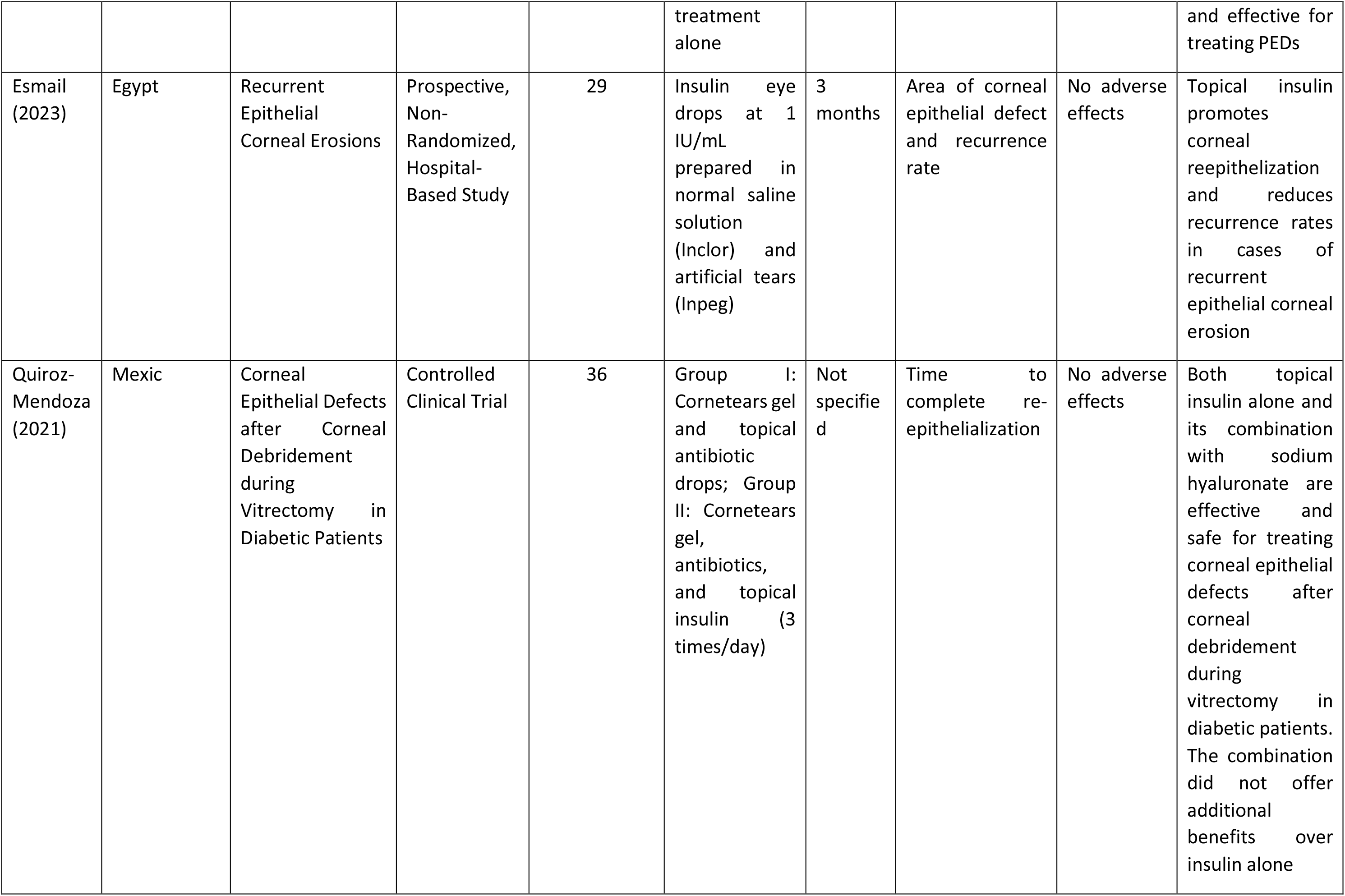
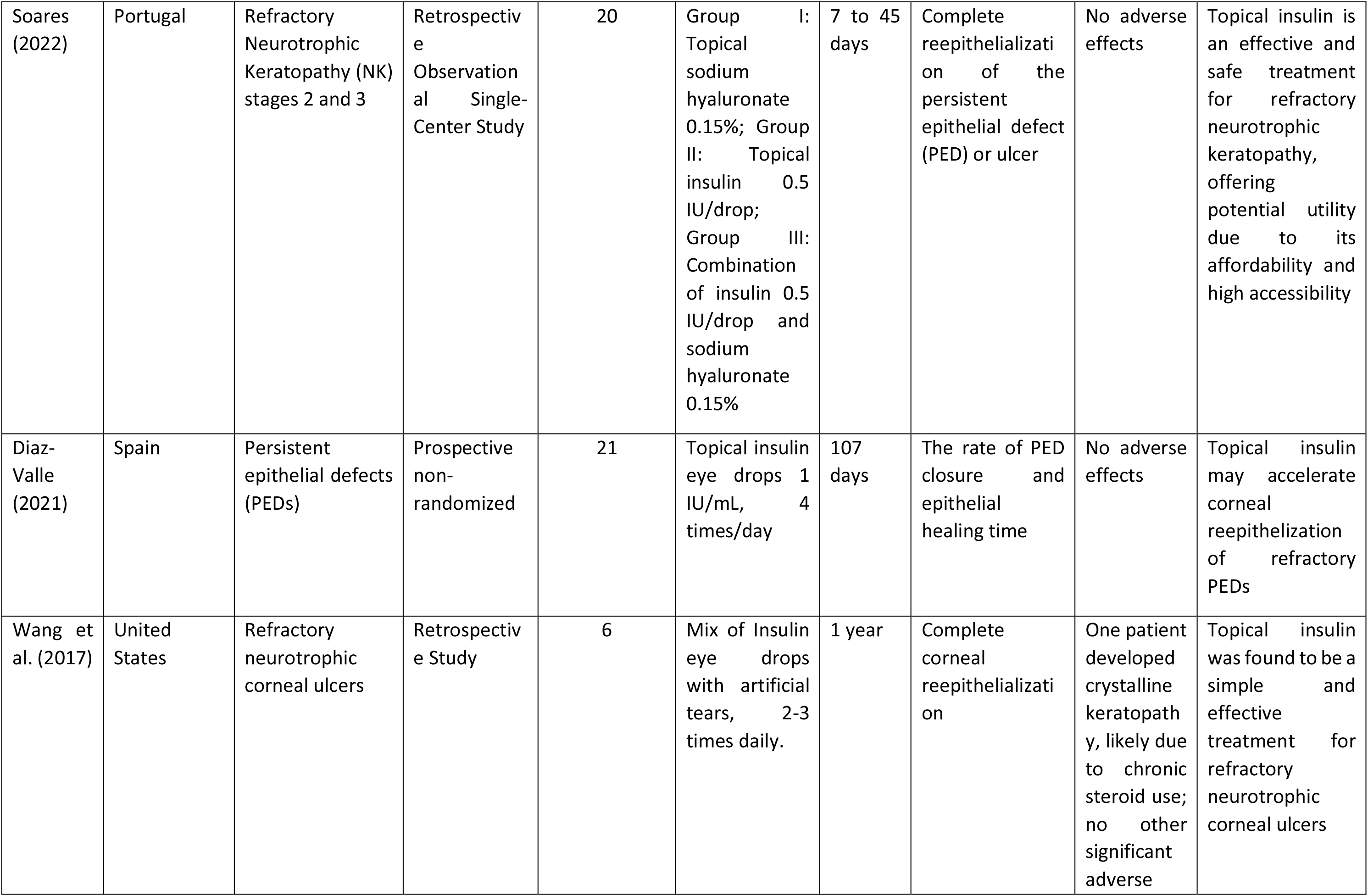
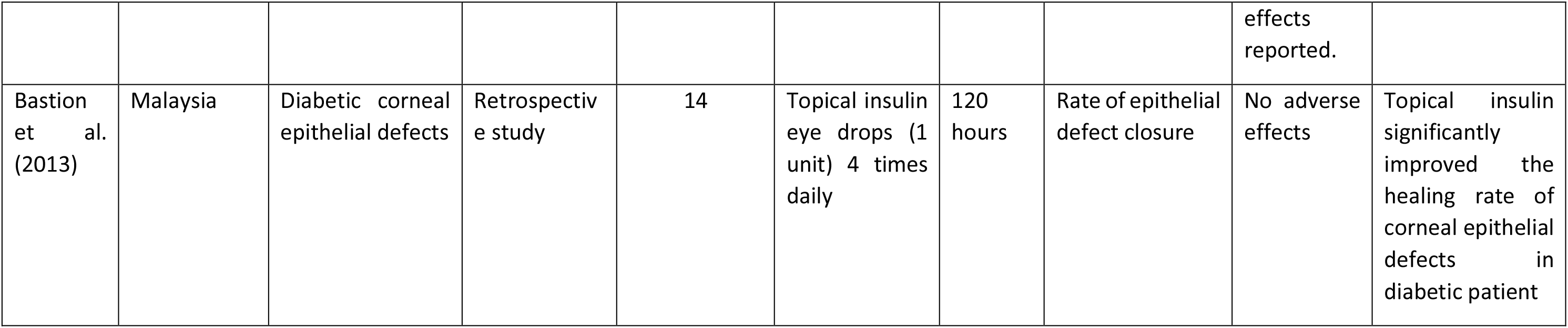
Key Studies on the Use of Topical Insulin Eye Drops for Various Ocular Pathologies.

Table 4 presents an overview of insulin eye drop formulations reported in clinical studies. This table summarizes key details, including insulin concentrations, sources, preparation processes, and administration practices.

**Tabel 4.** Summary of Insulin Eye Drop Formulations, Preparation Methods and Administration Practices in Clinical Studies.

| Authors & Publication | Insulin Concentration | Insulin Source | Preparation Process | Administration & Storage |
| --- | --- | --- | --- | --- |
| Fai et al. (Asia-Pacific Journal of Ophthalmology, 2017) | 0.5 units/drop,<br>1 unit/drop,<br>2 units/drop | Actrapid HM 100 U/mL (Novo Nordisk) | - 0.5 units/drop: 1.25 mL Actrapid (125 U) + 3.75 mL normal saline (25 U/mL).<br>- 1 unit/drop: 2.5 mL Actrapid (250 U) + 2.5 mL normal saline (50 U/mL).<br>- 2 units/drop: 5 mL Actrapid (500 U), no dilution. | Applied as 1 drop, 4 times daily. Best concentration: 0.5 units/drop. |
| Eleiwa et al. (Scientific Reports, 2024) | 1 unit/drop | Actarapid insulin (250 U) | 2.5 mL Actarapid (250 U) + 2.5 mL artificial tears (50 U/mL). | Applied as 1 drop, 4 times daily. Stored at 2°C. |
| Diaz-Valle et al. (Acta Ophthalmologica, 2022) | 1 IU/mL | Actarapid insulin (100 IU/mL) | Diluted in polyethylene glycol and polypropylene glycol base, sterile-filtered, and stored in amber glass bottles. | Applied as 1 drop, 4 times daily. Stored for up to 1 month. |
| Azmi & Bastion (Eye & Contact Lens, 2020) | 1 unit/drop | Actrapid HM 1000 U/mL (Novo Nordisk) | 1.25 mL Actrapid (1000 U/mL) + 3.75 mL sodium chloride 0.9% (25 U/mL). | Applied as 1 drop, 4 times daily. Stored in a sterilized 15-mL eyedropper bottle. |
| Abdi et al. (Scientific Reports, 2024) | 1 U/mL | Actrapid HM 1000 U (Novo Nordisk) | Diluted in Balanced Salt Solution (BSS) using sterile technique. | Applied as 1 drop, 4 times daily. Stored in a refrigerator. |
| Castro Mora et al. (Archivos de la Sociedad Española de Oftalmología, 2023) | 1 IU/mL to 100 IU/mL | Actrapid insulin (fast-acting) | Dissolved in artificial tears or polyethylene glycol and polypropylene glycol base. Prepared in a sterile environment. | Applied as 1 drop, 4-6 times daily, depending on the severity and type of pathology. |
| Soares et al. (Cornea, 2022) | 1 unit/mL | Fast-acting insulin | Diluted 1 unit of insulin in 1 mL of artificial tears with a propylene glycol base. | Applied as 1 drop, 4 times daily. Drops were preserved at 2°C. |
| Quiroz-Mendoza et al. (Revista Mexicana de Oftalmología, 2021) | 0.5 IU/drop | Humulin® R, Eli Lilly and Company, Indiana, USA | 1.25 mL of Humulin® R (100 IU/mL) was diluted with 3.75 mL of sterile 0.9% saline solution, resulting in a concentration of 25 IU/mL, equivalent to 0.5 IU/drop. | Applied as 1 drop, 4 times daily. The bottle was kept refrigerated and replaced if the treatment duration exceeded 3 days. |
| Vicario-de-la-Torre et al. (Pharmaceutics, 2024) | 1 IU/mL | Actrapid® 100 IU/mL (Novo Nordisk) | Two consecutive dilutions using isotonic sodium chloride solution (NaCl 0.9%) or artificial tears based on PEG and PG. Final sterile filtration before packaging. | Applied as 1 drop every 6 hours (four times daily). Stored in a refrigerator at 2–8°C. Shelf life estimated at 14-15 days. |
| Esmail et al. (Irish Journal of Medical Science, 2023) | 1 IU/mL | NovoRapid Penfill 100 units/mL (Novo Nordisk Limited Company, UK) | Regular insulin diluted in sodium chloride (SC) injection to achieve 1 IU/mL in a base of polyethylene and polypropylene glycol. Sterile technique used for preparation. | Applied as 1 drop every 6 hours (4 times daily). Stored in the refrigerator for up to one month. |
| Diaz-Valle et al. (European Journal of Ophthalmology, 2021) | 1 U/mL | Regular insulin in solution for subcutaneous injection | Diluted in a polyethylene glycol and polypropylene glycol base. Sterile filtration and packaging in sterile amber glass eye drop bottles. The drops were refrigerated and used up to 1 month after preparation. | Diluted in a polyethylene glycol and polypropylene glycol base. Sterile filtration and packaging in sterile amber glass eye drop bottles. The drops were refrigerated and used up to 1 month after preparation. |
| Wang et al. (Cornea 2017) | 1 unit/mL | Regular insulin in artificial tears | Diluted with artificial tears in a polyethylene glycol and propylene glycol base at a concentration of 1 unit per milliliter. | Applied as 2-3 drops daily. Stored in a refrigerator and used up to 1 month after preparation. |
| Bastion et al. (Med J Malaysia 2013) | 1 unit/drop | Actrapid HM 1000 U (Novo Nordisk) | 2.5 mL Actrapid (250 U) mixed with 2.5 mL normal saline to create a solution of 50 U/mL. Prepared using aseptic technique. | Applied as 1 drop 4 times daily. Stored in a commercial applicator bottle. Drops were discarded and a new preparation made every 3 days. |

### Discussion

#### Diabetic Keratophaty

Some studies provide a clear proof of the efficacy of topical insulin in accelerating the healing of the corneal epithelium with diabetic keratopathy mainly in post vitrectomy cases. The effect was documented in a range of concentration levels of insulin within the regimen: wound healing was faster and re-epithelization was better than in control eyes contributing to a new therapeutic strategy in the management of this complication.

One study demonstrated that diabetic patients treated with topical insulin experienced complete epithelial closure in an average of 60±15 hours, compared to 78±30 hours for those receiving conventional treatment. Notably, while no statistical difference was observed compared to non-diabetic patients (p>0.05), insulin treatment clearly accelerated healing in diabetic eyes without causing adverse effects (8). Further, dosage optimization appears crucial. In a randomized controlled trial, lower doses of insulin (0.5 units per drop) were found to be more effective than higher doses (1 and 2 units per drop), with a 100% healing rate observed in patients receiving the lower dose compared to 75% and 62.5% healing rates for the higher doses (p=0.036). This indicates that lower concentrations may enhance efficacy while minimizing the risk of side effects (9).

Other studies, including those by Eleika et al. and Dasrilsyah et al., reinforced the positive effects of insulin on corneal healing. Both reported faster epithelial healing rates in diabetic patients treated with insulin compared to those receiving artificial tears or conventional therapies (10,11). A retrospective case-control study was conducted by Eleika and colleagues with the purpose of assessing the effect of neurotrophic keratopathy within one month after diabetic vitrectomy among such patients who had been treated with preservative-free lubricating eye drops and prophylactic topical antibiotics compared to those patients receiving identical treatment but with one unit of insulin added per drop. The results indicated that insulin treatment significantly hastened the corneal epithelial healing by an average of 12.16 days as compared with the controls (p=0.001). Remarkably, no case in the insulin-treated group failed to achieve corneal epithelial healing in NK stage 2, while only 20% were failing in NK stage 3, as opposed to the control group that had a corresponding 20% and 66.7%, respectively (p<0.001). This study concludes that insulin effectively and safely enhances the healing of neurotrophic keratopathy after diabetic vitrectomy (10)Also Dasrilsyah et al. investigated the effect of 0.5U/ml insulin or artificial tears on postoperative corneal epithelial defects in patients with diabetes under vitrectomy for retinal detachment. Findings indicated that the healing rate at 36 h (p = 0.010), 48 h (P < 0.009) and after 144 h (p = 0.009) were significantly higher in insulin-treated group than artficial tears group respectively. In addition, the absolute rate of closure from baseline was significantly higher in the insulin-treated group (1.20 ± 0.29 mm²/h) compared with that in the artificial tears group (0.78 ± 0.20 mm²/h, p<0.001). This study also demonstrated that topical insulin has no negative systemic effects (11)In the treatment of corneal epithelial defects following pars plana vitrectomy, a matched-controlled clinical trial compared the efficacy of topical insulin with sodium hyaluronate and their combination. The study randomized 36 eyes into three groups: topical insulin (0.5 IU/drop), sodium hyaluronate (0.15%), and a combination of both. The results indicated that the insulin-treated group exhibited significantly faster re-epithelialization (3.00 ± 0.85 days) compared to the sodium hyaluronate group (4.25 ± 0.62 days) and the combined group (3.33 ± 0.98 days), with all p-values <0.001. Despite these differences in re-epithelialization time, the reduction in wound area was observed across all groups. However, the insulin group consistently showed a lower wound area at day one, although this was not statistically significant. Importantly, no treatment-related adverse effects were reported in any of the groups, confirming the safety of insulin and its potential as a standalone therapy. Notably, the combination of insulin with sodium hyaluronate did not provide any additional benefit over insulin monotherapy, suggesting that insulin alone may be sufficient to achieve optimal therapeutic outcomes in corneal epithelial healing (12)

#### Refractory persistent corneal epithelial defects and neurotrophic keratopathy

Several studies have explored the effectiveness of topical insulin eye drops in promoting healing for persistent epithelial defects (PEDs) and neurotrophic keratopathy (NK), with encouraging results across various patient populations.

In a prospective non-randomized study, Abdi et al. evaluated the impact of 1 U/mL insulin eye drops applied four times daily in 23 patients with refractory PEDs. The study showed significant improvement in 69.6% of the cases by day 50, with smaller epithelial defects (≤5.5 mm²) healing faster within 20 days. Patients with larger defects (>16 mm²) also showed substantial improvement, with 71% healing by day 50. (13)

Further supporting these findings, Diaz-Valle et al. compared the use of insulin eye drops against autologous serum for PEDs. In this study, 84% of patients treated with insulin achieved complete epithelialization, significantly higher than the 48% in the autologous serum group (p=0.002). Additionally, the time to epithelialization was shorter in the insulin group (32.6 ± 28.3 days) compared to the autologous serum group (82.6 ± 82.4 days), with lower recurrence rates and fewer surgical interventions required (p=0.011). This comparison highlights the superior efficacy of insulin over autologous serum, positioning it as a more reliable and efficient treatment option for PEDs.mThe requirement for amniotic membrane transplantation, AMT, was thus lesser in the insulin group, p=0.05, with a lower recurrence of PED because only 11% recurred in the insulin group against 43% in the autologous serum group, p=0.02. (14)

Vicario-de-la Torre et al. further investigated the stability and safety of two compounded insulin formulations (Inclor and Inpeg), demonstrating that Inclor, prepared with isotonic saline solution, was both more stable and effective, achieving complete epithelialization in all patients within a mean of 16.6 ± 10.8 days, with low recurrence rates and no adverse events. The formulation was well-tolerated, with viability above 90%, without adverse events reported. This suggests that certain insulin formulations may be optimized for better therapeutic outcomes (15).

Esmail et al. also observed significant improvements in recurrent epithelial erosion when insulin was combined with conventional PED management. Patients treated with insulin had reduced defect areas as early as two weeks after treatment initiation (p=0.006), with sustained improvements over two and three months (p=0.046 and p=0.002, respectively). Recurrence rates were lower in the insulin-treated group (11% vs. 21.4%), supporting the use of insulin for reducing recurrence in PEDs. The present study underlines, besides the efficacy of topical insulin in promoting corneal reepithelization, its advantages in reducing recurrence rates, excellent tolerance, availability, and cost-effectiveness (16)

In the context of neurotrophic keratopathy, insulin has shown similar promise. A study by Wang et al. assessed six patients with recalcitrant neurotrophic corneal ulcers, achieving complete re-epithelialization in all cases within 7 to 25 days. In the retrospective series reported the results of topical insulin, 1 unit/mL, administered four times daily in the management of stages 2 and 3 refractory NK. Nine patients with NK of different causes were included, which had developed after herpetic keratitis, following penetrating keratoplasty, in diabetes mellitus, acoustic neuroma resection, gunshot injury to the head, and abusive use of topical nonsteroidal anti-inflammatory medications. The corneal appearance, before and after insulin treatment, was photographed both with and without cobalt-blue filter and fluorescein stain. The days until complete reepithelialization were counted in each case. Results demonstrated that full reepithelialization was achieved in 90%. Importantly, the average time to reepithelialization was significantly shorter in NK stage 2 compared with NK stage 3: 18 ± 9 days versus 29 ± 11 days, p=0.025. This reinforces the effect of topical insulin in stimulating the healing of the cornea, especially in NK with minimal damage. Improvement in BCVA was also significant in NK stage 2 patients (p<0.001) and stage 3 patients (p=0.004), further supporting the therapeutic potential of insulin eye drops. Of note, no adverse effects were reported during the follow-up, confirming the safety and tolerance of this therapeutic approach. (17)

Diaz-Valle et al. conducted a prospective study involving 21 patients with PEDs resistant to medical treatments, reporting an encouraging 81% of treated eyes achieving complete epithelialization within an average of 34.8 ± 29.9 days. Even in cases where full healing was not achieved, the size of epithelial defects was significantly reduced by an average of 91.5%. These findings suggest that insulin is a valuable and safe alternative for managing difficult cases of PEDs, particularly when standard therapies are ineffective (18).

In a separate retrospective study, Wang et al. investigated the use of insulin eye drops for recalcitrant neurotrophic corneal ulcers in six patients who had previously failed other medical and surgical treatments. Complete re-epithelialization was achieved in all subjects within 7 to 25 days, highlighting the efficacy of insulin in promoting corneal healing in these resistant cases. The treatment was well-tolerated, with only one patient developing crystalline keratopathy, likely due to chronic steroid use and not directly related to the insulin treatment. These findings reinforce the potential of insulin as a simple, effective, and safe therapy for neurotrophic corneal ulcers, warranting further investigation for broader clinical applications(19)

Taken together, these studies demonstrate that topical insulin is a safe, cost-effective, and promising therapeutic agent for promoting corneal healing in patients with refractory PEDs and neurotrophic keratopathy, outperforming conventional treatments like autologous serum and artificial tears in terms of healing time and recurrence rates.

#### Dry eye

Therefore, this comparison between topical insulin and standard artificial tears regarding the treatment of dry eye disease in diabetic patients can give some insight into a comparison of their use and what potential benefits may be obtained with insulin therapy. In the investigation of Azmi et al., 80 eyes were treated for four weeks with 1 unit of insulin eye drops four times daily; meanwhile, another 80 eyes received standard artificial tears. Following four weeks of baseline, the measurements-the Ocular Surface Disease Index (OSDI), Schirmer test 1, tear break-up time, and the results from the Ocular Sjögren’s International Collaborative Clinical Alliance (SICCA)-were obtained. Significant improvement in OSDI scores was established with an improvement in 66% of the insulin treatment group and 63% of the artificial tears treatment group (p=0.001). These findings indicate that, although both treatments are effective, eye drops of insulin could provide at least an alternative to artificial tears in the standard management of symptoms of dry eye in diabetic patients. (20)

In a more recent prospective study, designed to further support the potential of insulin therapy in dry eye disease, the inclusion comprised 32 eyes with persistent signs and symptoms of dry eye despite intensive lubricating treatments and prior use of autologous serum for at least six months (71%), as well as cyclosporine (63%). In this study, insulin eye drops were introduced on a compassionate basis as an adjunct to existing treatment. The mean symptom score at baseline was 3.4 ±1.3 out of 5, mean hyperemia score was 1.0 ± 0.9, and the corneal staining score was 2.5 ±1.3 for these patients. After three months of insulin treatment, symptom scores had significantly improved to 2.3±1.0, p=0.001, with significant decrease in hyperemia score, 0.3±0.4, and corneal staining score, 1.1±1.0, both with a p < 0.001. Subjective assessments also showed that 31% felt much better, 38% better, 19% slightly better, whereas only 13% reported no change. This is further confirmed by the absence of adverse events, including infection, which strengthens insulin eye drops as an adjunctive treatment for dry eye disease (21)

These findings collectively suggest that topical insulin may offer a valuable therapeutic option for managing dry eye disease, particularly in cases where standard treatments have failed. The improvements in both objective measures and patient-reported outcomes indicate that insulin eye drops not only alleviate symptoms but also enhance the overall health of the ocular surface.

## Conclusion

To this effect, the conclusions from our systematic review resulted in strong evidence in support of topical insulin as a potential therapeutic agent for different ocular surface disorders. It had a promising role in improving corneal healing and managing refractory conditions. These findings also suggested that such patients with non-healing, persistent epithelial defects, diabetic keratopathy, and other problem ocular conditions would definitely benefit from adding topical insulin to their treatment modalities. We believe that this work will drive further investigation in the promising therapy, underlining opportunities that can be gained by the use of insulin in order to enhance patient outcomes in ophthalmic care. Larger and well-designed clinical trials need to be conducted to uniformly develop treatment protocols for full clinical validation toward broader applications of topical insulin.

## Author contributions: CRediT

DP: Supervision, Project administration, Writing – review & editing. RBR: Conceptualization, Investigation, Methodology, Project administration, Resources, Writing – original draft. SF: Conceptualization, Methodology, Supervision, Project administration, Resources, Validation, Writing – review & editing.

## Data Availability

All data produced in the present work are contained in the manuscript

